# Divergent climate suitability profiles for dengue and chikungunya transmission by Aedes albopictus in Mauritius

**DOI:** 10.64898/2026.08.23.26361159

**Authors:** Mohabeer Teeluck, Emma S. McBryde, Oyelola A. Adegboye, Stephan Karl, Benn Sartorius, Eloise B. Skinner

**Author notes:** Corresponding author: Mohabeer Teeluck.

## Abstract

**Background:** Empirical surveillance for *Aedes*-borne arboviruses is inherently reactive, detecting transmission after it has commenced. For small island settings where dengue and chikungunya circulate sporadically, characterising when and where environmental conditions could support local transmission is critical for preparedness. In Mauritius, *Aedes albopictus* is the sole primary vector for dengue and chikungunya viruses, but previous suitability assessments have relied on *Aedes aegypti* parameterisation.

**Methods:** We estimated monthly Index P for dengue and chikungunya across 160 localities in Mauritius from January 2014 to October 2024. Index P, a mechanistic transmission suitability measure derived from the Ross-Macdonald framework that climate-dependent transmission potential attributable to one adult female mosquito. Mean temperature and relative humidity were derived from ERA5-Land reanalysis dataset via Google Earth Engine and incorporated within the Mosquito-borne Viral Suitability Estimator (MVSE) framework. Index P was also parameterised with *Ae. albopictus*-specific biological priors and virus-specific vector competence values for both dengue and chikungunya.

**Results:** Transmission suitability for both viruses was concentrated within the austral summer (November to April), with near-zero values in winter, below the indicative transmission threshold (Index P ≥ 0.5). Chikungunya exhibited consistently higher, more spatially widespread and longer-lasting suitability than dengue: all districts exceeded the transmission suitability threshold for chikungunya (Index P = 0.71), while median dengue Index P = 0.24, remaining below this threshold, during the same study period. Dengue peak suitability was concentrated in western coastal localities, consistent with the greater thermal sensitivity of its extrinsic incubation period in *Ae. albopictus*.

**Conclusions:** These findings indicate that dengue and chikungunya have distinct, virus-specific climate-suitability profiles in Mauritius, and should not be treated as interchangeable for preparedness purposes. This provides an important *Ae. albopictus*-parameterised evidence base for Mauritius, enabling seasonal and geographic targeting of surveillance and vector control ahead of, rather than in response to local transmission.

## Introduction

Tropical and subtropical islands are recurrent hotspots for *Aedes*-borne arbovirus outbreaks, with dengue virus (DENV), chikungunya virus (CHIKV) and Zika virus causing successive epidemics across island nations in the Indian Ocean, the Pacific and the Caribbean (1,2). High international connectivity can facilitate repeated pathogen introductions into immunologically naive populations, while in island settings with limited health-system capacity, delayed detection and constrained resources may hinder response (3–5). However, islands differ greatly in connectivity, immunity profiles and surveillance/health-system capability. Globally, dengue places nearly half the world’s population at risk, with as many as 100-400 million infections annually (6), while chikungunya outbreaks have expanded in frequency and geographic reach, driven partly by viral adaptation favouring *Aedes albopictus*-mediated transmission and by introduction into populations with low pre-existing immunity (7). The southwestern Indian Ocean (SWIO) islands exemplify these dynamics: repeated arbovirus introductions, established *Aedes* vector populations and suitable climatic conditions have sustained recurrent dengue and chikungunya outbreaks across the Mascarene archipelago and neighbouring island states (8,9).

Climate shapes *Aedes*-borne virus transmission through strongly non-linear effects on mosquito life-history traits and viral development. Temperature regulates larval development rate, adult longevity, biting frequency, extrinsic incubation period (EIP) and vector competence, with transmission suitability generally peaking at intermediate temperatures and declining sharply under thermal extremes (10–14). Relative humidity modulates adult mosquito survival through desiccation stress, with higher humidity extending longevity and sustaining vector activity (15,16). Together, temperature and humidity define the environmental window within which local arbovirus circulation is biologically feasible, and this window varies in duration, timing and intensity across geographic settings. For island settings where outbreaks are sporadic rather than sustained, understanding when and where climatic conditions permit local transmission is essential for preparedness.

Mauritius provides a unique case for examining climate-driven arbovirus transmission suitability. The island is non-endemic for both dengue and chikungunya but has experienced recurrent outbreaks following importation events, including sporadic dengue transmission since 2009, an unprecedented surge exceeding 7,000 dengue cases in 2024, and a major chikungunya epidemic in 2005–2006 (17–19). Crucially*, Ae. albopictus* is the sole established primary vector for both viruses in Mauritius, with *Ae. aegypti* thought to have been absent since the late 1940s (20). Its life-history traits, including adult survival, biting behaviour and EIP, are strongly temperature- and humidity-dependent (13,15,21,22) and the island’s volcanic topography, rising from warm coastal lowlands to a cooler central plateau creates pronounced environmental gradients that may generate spatial heterogeneity in transmission suitability across a relatively small land area (25).

Existing software/tools such as ARBOCARTO/SIT and ALBOMAURICE predict *Ae. albopictus* abundance and distribution using deterministic temperature- and rainfall-dependent traits (21,26–28) but do not incorporate virus-specific transmission parameters. A systematic review of climate and mosquito-borne disease relationships in small island and developing states, including Mauritius, confirmed this evidence gap (29). Mechanistic climate-suitability frameworks such as the Mosquito-borne Viral Suitability Estimator (MVSE) (30) address this directly by integrating temperature- and humidity-sensitive vector, virus and human biological parameters into an estimate of transmission suitability (Index P), derived from the Ross–Macdonald transmission framework (31). MVSE has been applied to estimate dengue suitability globally and across regional contexts (30,32–34), including to Mauritius using *Ae. aegypti* parameterisation (9). However, to our knowledge, no published study has estimated Index P for Mauritius using *Ae. albopictus*-specific biological priors, despite this being the sole primary vector on the island. This study therefore aims to provide an *Ae. albopictus*-parameterised application of Index P for dengue and chikungunya in Mauritius. Specifically, our objectives are to 1) characterise the temporal structure and duration of transmission suitability windows across the island; 2) identify sub-national spatial heterogeneity in suitability driven by the island’s climatic gradients; and 3) compare suitability profiles between dengue and chikungunya to assess whether the two viruses differ in their climate sensitivity within this system.

## Methodology

The Republic of Mauritius is a Small Island Developing State (35) located in the southwestern Indian Ocean, comprising the main island (approximately 1,868 km^2^) and smaller islands dependencies. The main island of Mauritius is subdivided into nine districts (36) and, more finely 160 localities (Figure 1). The island’s volcanic topography rises progressively from low coastal plains to a central highland plateau (Figure 1), creating distinct environmental gradient between coastal and inland areas (25). Mauritius has a tropical maritime climate, with warm, humid summers (November-April; mean 24.7 °C) and cooler, drier winters (May-October; mean 20.4 °C), and considerable spatial climatic variation shaped by elevation and the prevailing southeast trade winds (37–39) Full district-level climate summary statistics are provided in Supplementary Table S1.

**Figure 1:**
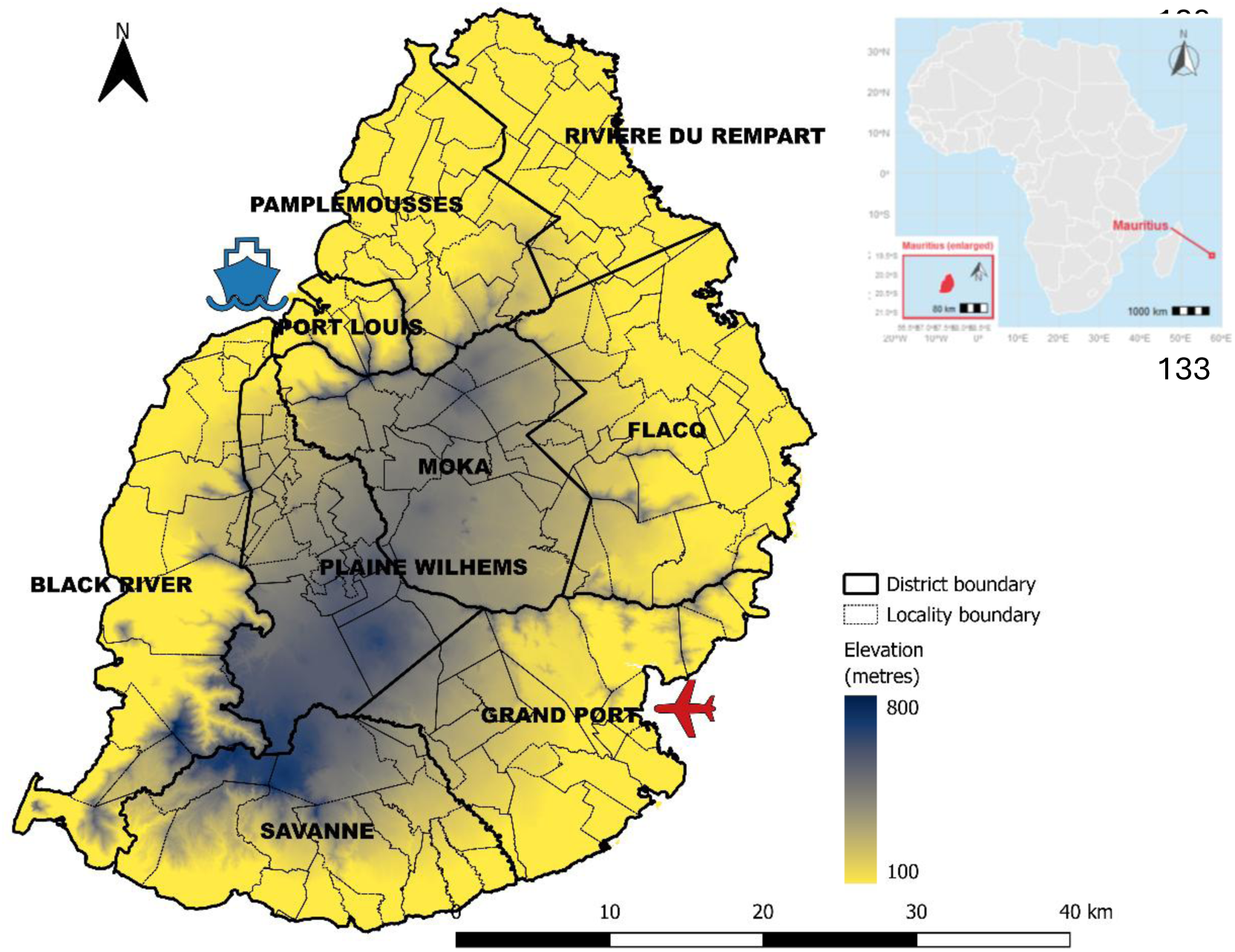
Topographic gradient and administrative boundaries of mainland Mauritius. The map illustrates the spatial variation in elevation across mainland Mauritius, with lower coastal areas surrounding a higher central plateau. District boundaries show the island’s principal administrative divisions, whereas locality boundaries highlight finer-scale study areas (towns and villages).

Daily mean temperature and dewpoint temperature were extracted at the locality level from ERA5-Land reanalysis datasets via Google Earth Engine (40,41) and aggregated to monthly means for December 2013-October 2024. Full extraction methods, including the zonal reduction and buffering procedure used to ensure complete locality coverage, are provided in Supplementary Methods S1. Relative humidity was derived from monthly mean temperature and dewpoint temperature using the Tetens/Clausius-Clapeyron approximation as shown in Supplementary Methods S2 (42,43).

Index P, a climate-driven measure of arbovirus transmission suitability, was estimated using the MVSE package in R (30). Index P is derived from a mechanistic expression of the basic reproduction number (R_0_) based on the Ross-Macdonald framework (31), integrating biting rate, adult mosquito lifespan, extrinsic and intrinsic incubation periods, human infectious period, and transmission probabilities (30) as shown in Equation 1 and Equation 2, in Supplementary Methods S3. It is interpreted as the average number of new human infections generated by a single infected *Ae. albopictus* female under given climate conditions, in a fully susceptible population, with values of Index P ≥0.5 considered indicative, though not definitive, of elevated climatic suitability for transmission (34).

We adapted this framework which was originally developed for *Ae. aegypti-*mediated dengue transmission (30,34), using *Ae. albopictus-*specific biological priors. Where possible, parameter estimates were drawn from Reunion Island, a geographically and ecologically proximate setting to Mauritius, including adult mosquito lifespan, the extrinsic incubation period for CHIKV, and the intrinsic incubation period for CHIKV (22,45,46). Where local or regional *Ae. albopictus*-specific parameters were unavailable, priors for the intrinsic incubation period for DENV and the human infectious periods for both viruses were drawn from published empirical studies (47–51). Biting rate and the extrinsic incubation period for DENV were derived from deterministic temperature-dependent Briere equations (13), while relative humidity was incorporated through its effect on adult mosquito survival (46). Full parameter definitions, prior distributions and sources are provided in Supplementary Table S2. Index P was further scaled using virus-specific vector competence coefficients as transmission efficiency multipliers for dengue and chikungunya (22,52). Biological parameter uncertainty was propagated through repeated sampling from prior distributions, with the 2.5th and 97.5th percentiles of the resulting Index P distributions used as uncertainty bounds (53). Additional details of the Index P equations, virus-specific vector competence adjustment, uncertainty propagation, and implementation are provided in Supplementary Methods S3.

## Results

Mean temperature across Mauritius was approximately 23.5 °C nationally, ranging from 22.9 °C in Plaines Wilhems to 24.1 °C in Riviere du Rempart, with coastal districts consistently warmer than the central plateau (Supplementary Table S3). Clear seasonal variation was evident, with the warmest months from January to March and declining temperatures from April onward (Supplementary Figure S1). National mean relative humidity was estimated to be 75.5%, peaking in April before declining sharply in early winter; the western region, particularly Black River, was comparatively drier (Supplementary Figure S2).

Nationally, Mauritius exhibited higher baseline transmission suitability and greater variability for chikungunya (median Index P = 0.71; 95% uncertainty interval (UI): 0.13-2.1), than dengue (median Index P = 0.32; 95% Ul: 0.03-0.8), although the overlapping uncertainty intervals indicate this difference should be interpreted cautiously (Table 1; Supplementary Figure S3). District-level estimates varied spatially for both viruses: All districts exhibited median chikungunya Index P values above the indicative transmission threshold (Index P ≥ 0.5), with lower values in the west and on the central plateau, whereas dengue remained below this threshold across all districts, indicating more uniformly low suitability. Warmer northern coastal districts (Rivere du Rempart and Pamplemousses) had the highest chikungunya suitability, while the more humid eastern districts (Grand Port, Flacq and Rivere du Rempart) showed relatively greater, though still sub-threshold, dengue suitability.

**Table 1.** Descriptive summary of Climate-Driven Transmission Suitability (Index P) of Chikungunya and Dengue at National and Districts level.

| Admin level |  | Chikungunya |  |  | Dengue |  |
| --- | --- | --- | --- | --- | --- | --- |
| National | Mean (SD) | Median (Range) | 95%<br>Uncertainty<br>Interval (UI) | Mean (SD) | Median (Range) | 95%<br>Uncertainty<br>Interval (UI) |
| Mauritius | 2.98 (4.46) | 0.71 (0.01-30.87) | 0.13 -2.1 | 1.18 (1.86) | 0.24 (0-18.05) | 0.03-0.8 |
| District | Mean (SD) | Median (Range) | 95%<br>Uncertainty<br>Interval (UI) | Mean (SD) | Median (Range) | 95%<br>Uncertainty<br>Interval (UI) |
| Riviere du Rempart | 3.25 (4.75) | 0.82 (0.02-25.45) | 0.15-2.35 | 1.26 (1.93) | 0.26 (0-11.13) | 0.03-0.83 |
| Pamplemousses | 3.31 (4.86) | 0.82 (0.02-23.51) | 0.15-2.32 | 1.29 (2.06) | 0.23 (0-12.53) | 0.02-0.8 |
| Grand Port | 2.65 (3.53) | 0.76 (0.01-17.99) | 0.13-2.19 | 1.04 (1.41) | 0.28 (0-7.72) | 0.04-0.83 |
| Flacq | 2.9 (4.04) | 0.73 (0.01-22.03) | 0.15-2.23 | 1.13 (1.63) | 0.26 (0-11.06) | 0.04-0.86 |
| Savanne | 2.98 (4.41) | 0.72 (0.01-24.87) | 0.12-2.11 | 1.23 (1.9) | 0.25 (0-12.44) | 0.04-0.83 |
| Port Louis | 3.3 (5.14) | 0.69 (0.02-24.12) | 0.12-2.09 | 1.29 (2.08) | 0.24 (0-11.54) | 0.03-0.8 |
| Black River | 3.18 (5.39) | 0.65 (0.01-30.87) | 0.12-2.14 | 1.37 (2.54) | 0.23 (0-18.05) | 0.03-0.82 |
| Moka | 2.67 (3.94) | 0.61 (0.01-24.12) | 0.11-1.9 | 1.15 (1.82) | 0.2 (0-9.65) | 0.02-0.7 |
| Plaine Wilhems | 2.86 (4.58) | 0.58 (0.01-24.12) | 0.11-1.83 | 1.03 (1.69) | 0.19 (0-10.75) | 0.02-0.7 |

A consistent seasonal pattern was observed for both viruses, with suitability peaking during the austral summer (November-April) and declining to near-zero during winter (May-October) (Figure 2). Chikungunya exhibited consistently higher vector-competence-adjusted Index P values than dengue throughout the study period, with the most pronounced during peak suitability months and narrowing during low-suitability winter months. The interquartile range remained constrained during periods of low suitability but broadened during peak seasons. Transmission-window duration (months with Index P ≥0.5) fluctuated across the study period: dengue windows were relatively consistent, lasting approximately 4-7 months, with the most prolonged period recorded in 2023, while chikungunya windows were more variable, ranging from approximately 4-9 months. Virus-specific differences in window duration were most evident in 2017, whereas 2023 was characterised by extended suitability for both viruses (Supplementary Figure S4).

**Figure 2:**
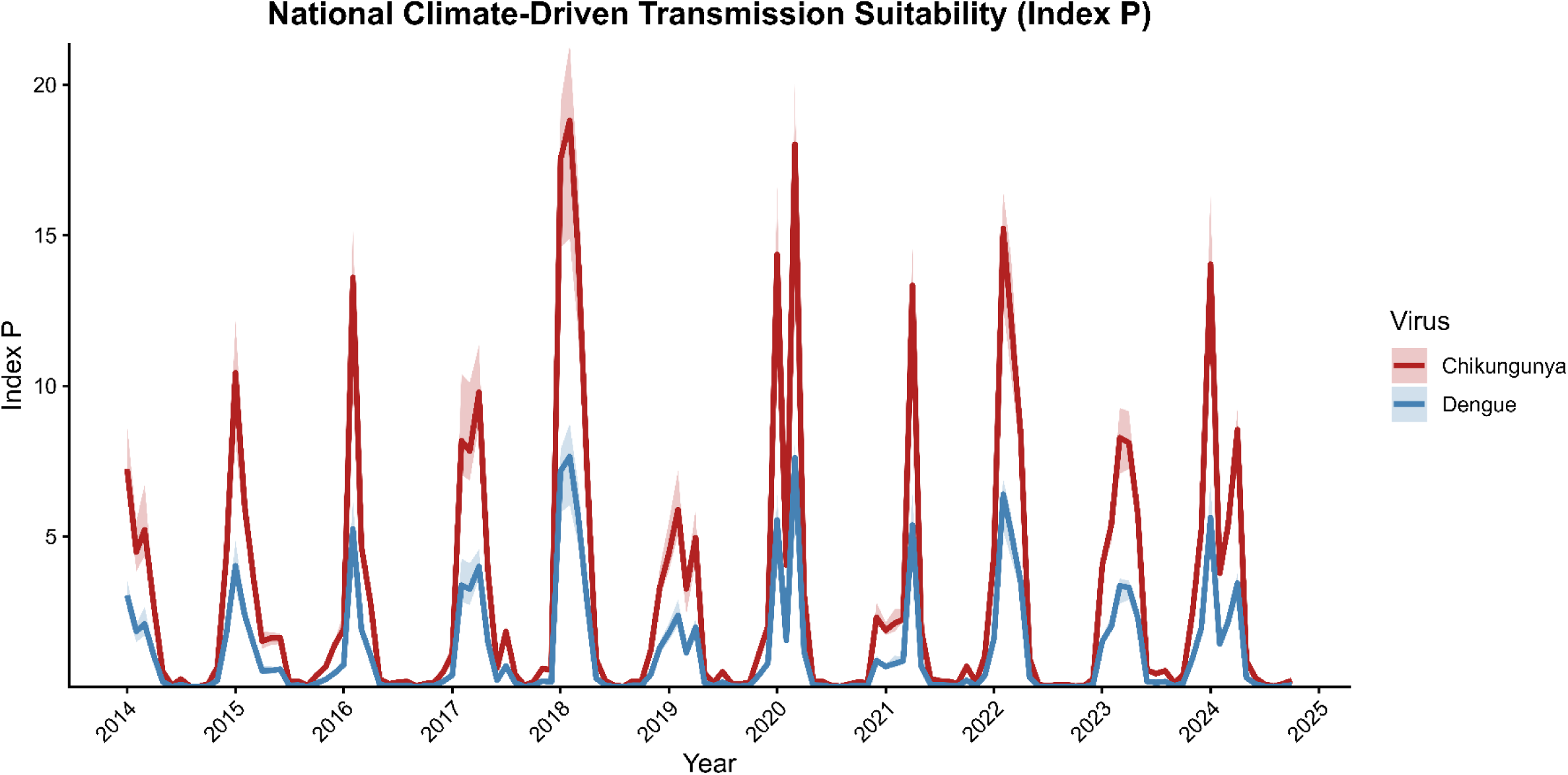
National monthly time series of Index P for DENV and CHIKV transmission suitability in Mauritius, 2014–2024. The figure demonstrates the temporal variation in Index P estimates, with higher values indicating more favourable conditions for virus-specific transmission by Ae. albopictus mosquitoes.

Peak transmission suitability (95th percentile of Index P) for both viruses was concentrated in coastal localities, though spatial patterns differed (Figures 3A and 3B). Chikungunya showed broadly distributed high suitability across coastal regions, particularly in the west, north, and to a lesser extent in the east and south. Dengue peak suitability was more geographically constrained, concentrated primarily in western localities within Black River district, which consistently exhibited the highest suitability for both viruses under favourable conditions.

**Figure 3A:**
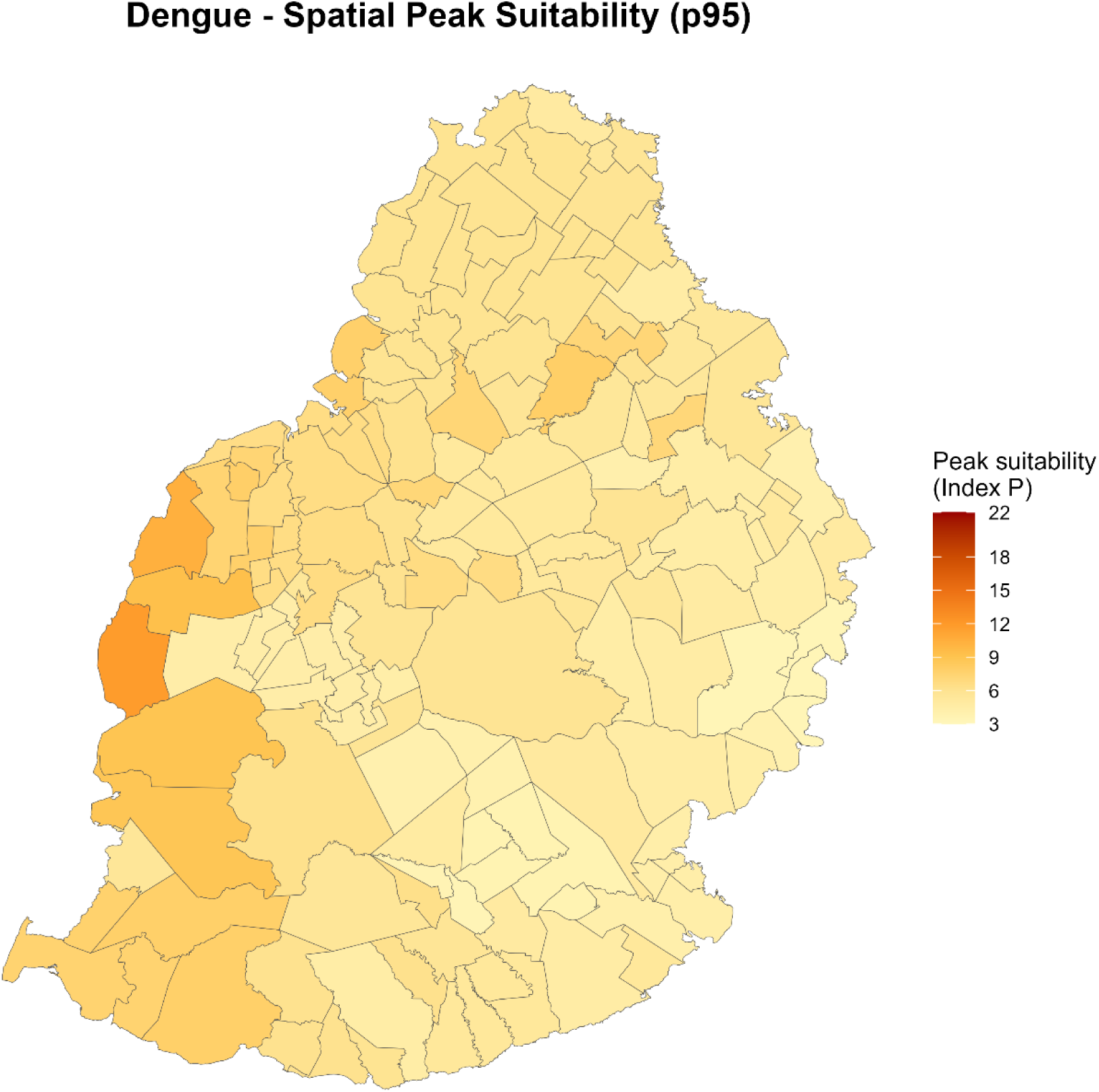
Spatial distribution of peak Index P suitability for dengue in Mauritius, 2014–2024. Maps show the estimated peak Index P value for each locality across the study period. Darker colours indicate higher peak climate-driven suitability for Ae. albopictus-mediated transmission. Supplementary Table S4 provide the corresponding peak Index P estimates for all localities.

**Figure 3B:**
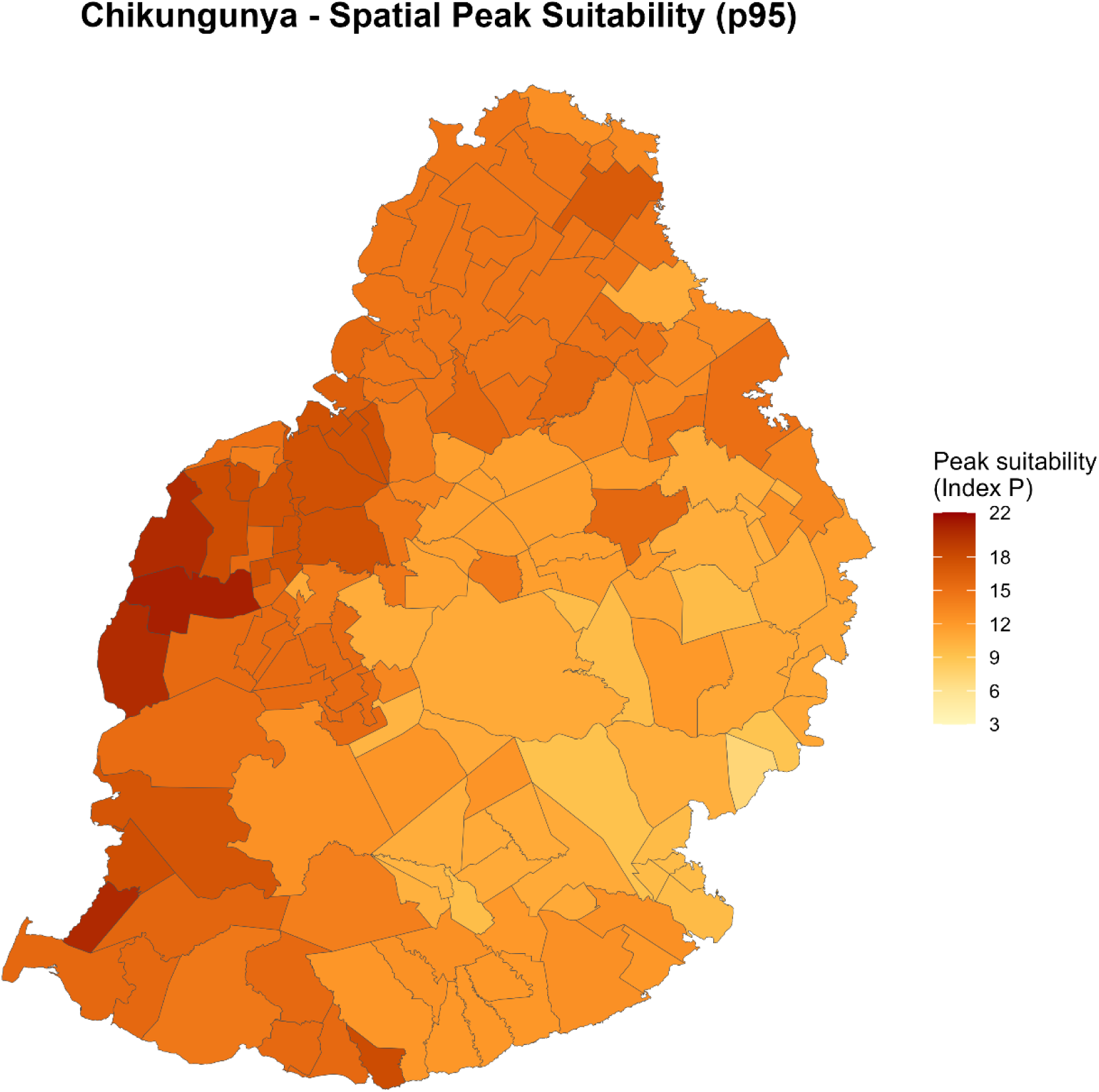
Spatial distribution of peak Index P suitability for chikungunya in Mauritius, 2014–2024. Maps show the estimated peak Index P value for each spatial unit across the study period. Darker colours indicate higher peak climate-driven suitability for Ae. albopictus-mediated transmission. Supplementary Table S4 provide the corresponding peak Index P estimates for all localities.

Transmission suitability for both viruses exhibited a clear seasonal spatial progression, remaining low across the island during winter and increasing gradually from November onward (Figures 4-5). For chikungunya, the November-December period marked the onset of elevated suitability. This initially concentrated along the northern and eastern coasts before spreading to broader coastal regions in January, extending further inland through February, and achieving near-uniform island-wide coverage by April with the highest values in northwestern districts (Figure 4A); suitability then declined from May onwards (Figure 4B). For dengue, coastal districts, particularly in the north and east, showed slightly elevated Index P values (though remaining below the threshold) during November-December, while the central plateau (Moka and Plaines Wilhems) remained lower (Figure 5A). Suitability increased gradually from January and expanded inland, reaching a more uniform distribution by April despite declining temperatures (Figure 5A), before declining more sharply than chikungunya from May onward (Figure 5B). Overall, dengue suitability was more spatially variable, while chikungunya remained consistently elevated and geographically widespread.

**Figure 4A:**
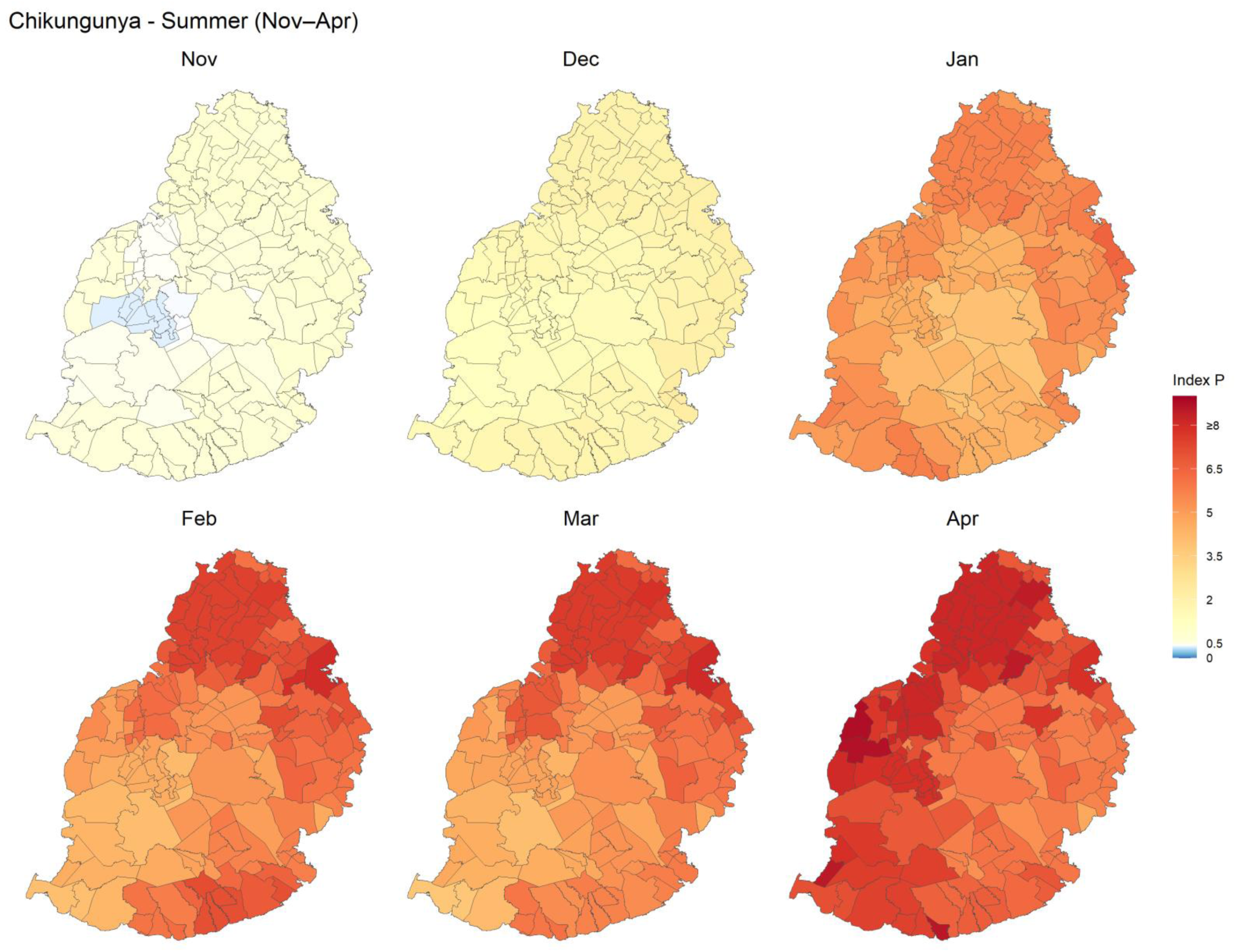
Seasonal spatial patterns of Index P median transmission suitability for chikungunya during summer in Mauritius, 2014–2024. Index P values above 0.5 indicate suitable conditions for transmission, with warmer colours representing higher climate-driven suitability for Ae. albopictus-mediated chikungunya transmission. Corresponding locality-level Index P estimates are provided in Supplementary Table S5.

**Figure 4B:**
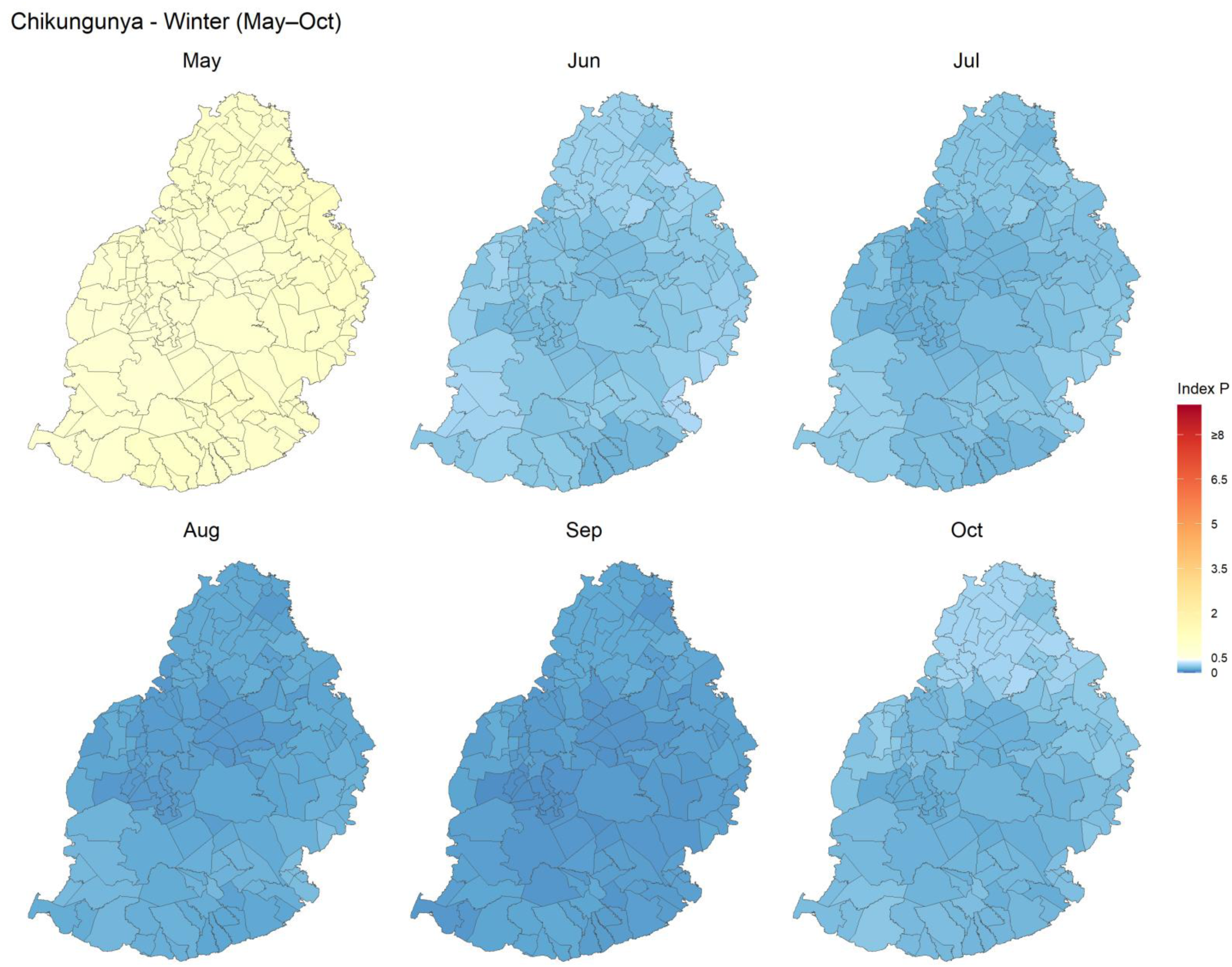
Seasonal spatial patterns of Index P median transmission suitability for chikungunya during winter in Mauritius, 2014–2024. Index P values above 0.5 indicate suitable conditions for transmission, with warmer colours representing higher climate-driven suitability for Ae.s albopictus-mediated chikungunya transmission. Corresponding locality-level Index P estimates are provided in Supplementary Table S5.

**Figure 5A:**
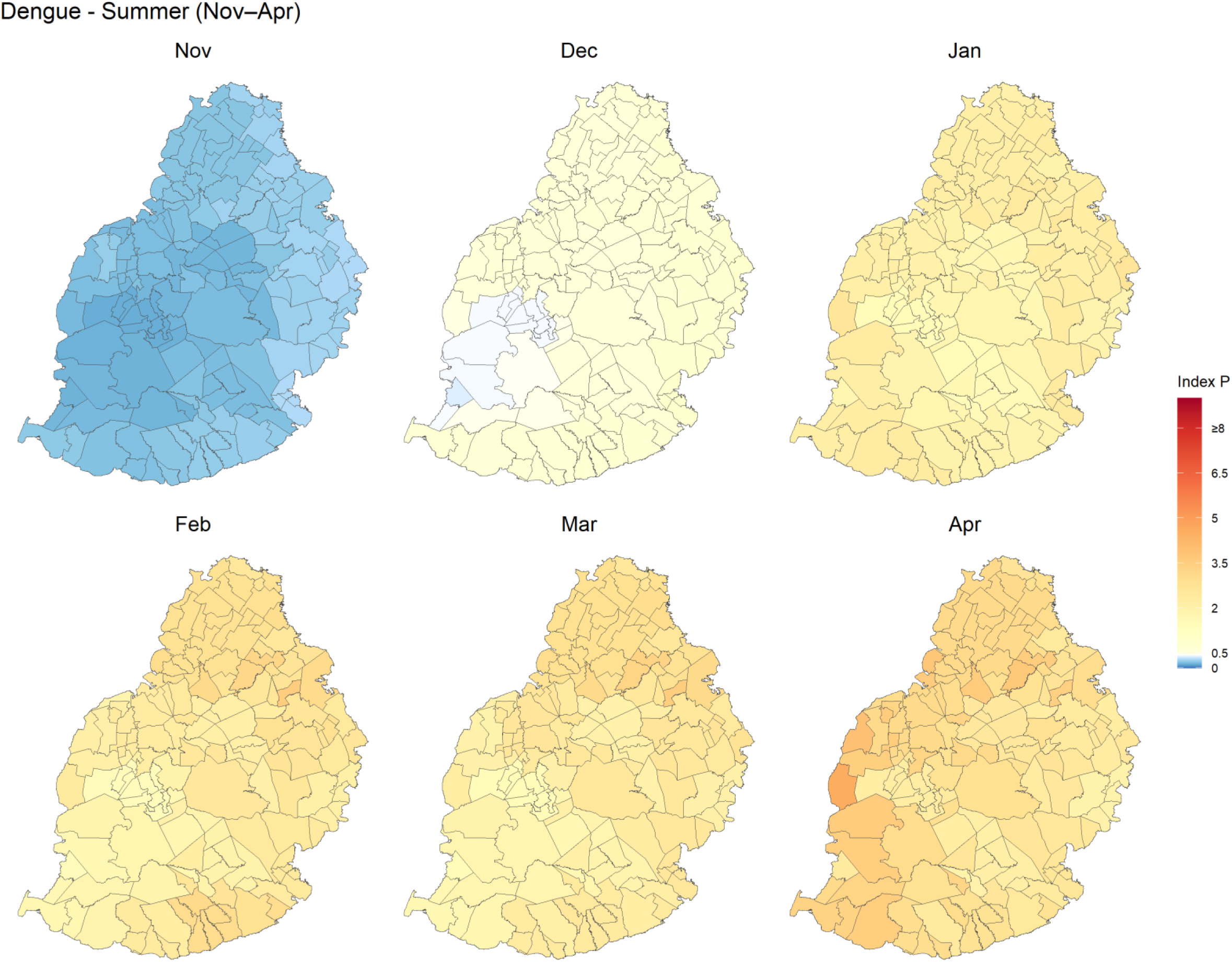
Seasonal spatial patterns of Index P median transmission suitability for dengue during summer in Mauritius, 2014–2024. Index P values above 0.5 indicate suitable conditions for transmission, with warmer colours representing higher climate-driven suitability for Ae. albopictus-mediated chikungunya transmission. Corresponding locality-level Index P estimates are provided in the Supplementary Tables.

**Figure 5B:**
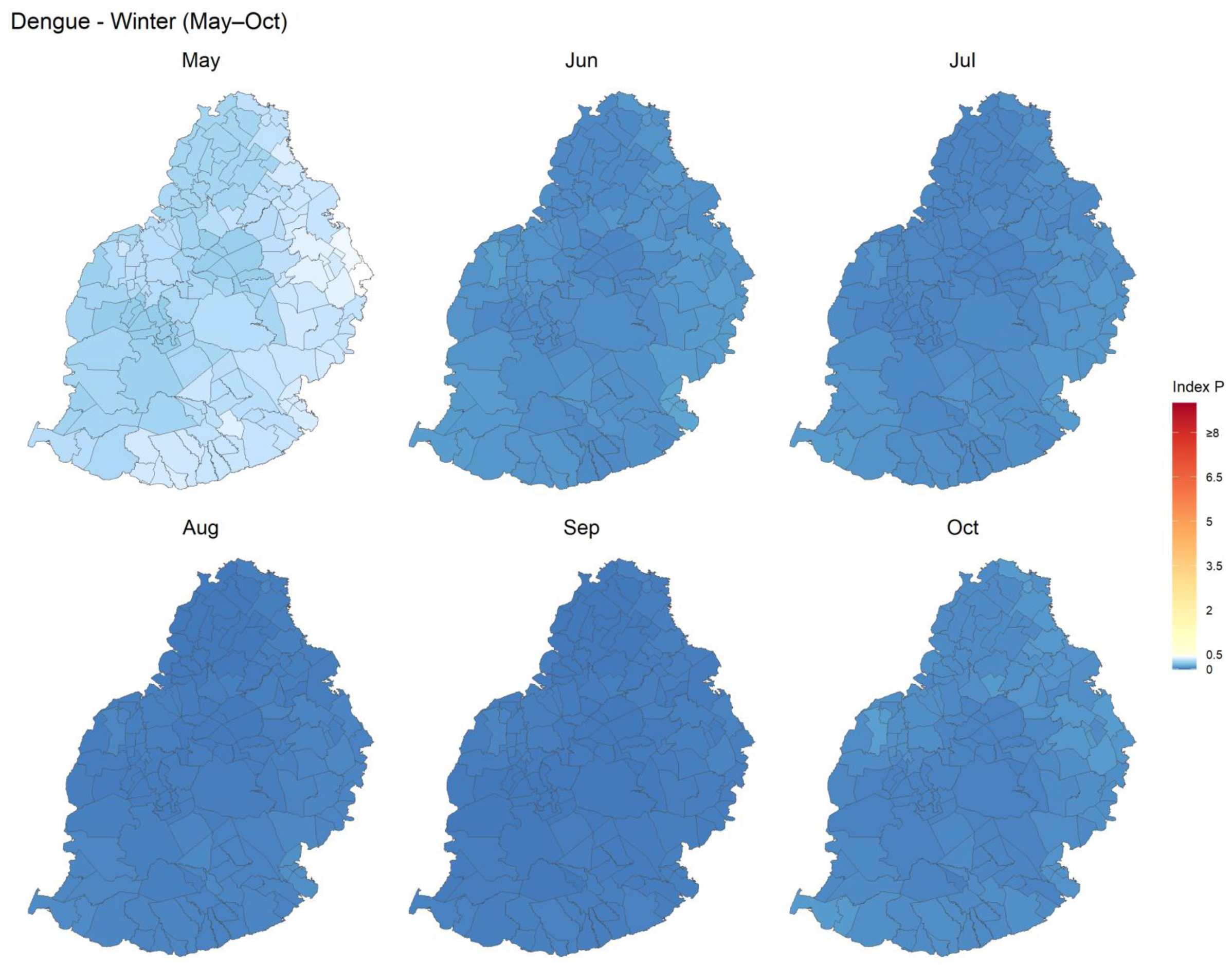
Seasonal spatial patterns of Index P median transmission suitability for dengue during winter in Mauritius, 2014–2024. Index P values above 0.5 indicate suitable conditions for transmission, with warmer colours representing higher climate-driven suitability for Ae. albopictus-mediated chikungunya transmission. Corresponding locality-level Index P estimates are provided in the Supplementary Tables.

## Discussion

This study estimates the seasonal and spatial patterns of climate-driven transmission suitability for dengue and chikungunya by *Ae. albopictus* in Mauritius using the MVSE Index P framework (30). Because *Ae. albopictus* is the sole established vector in Mauritius, vector-specific parameterisation is essential for generating suitability estimates relevant to local transmission ecology, and by incorporating virus-specific vector competence as a transmission efficiency multiplier, this study differentiates suitability profiles for dengue and chikungunya.

Transmission suitability for both viruses was concentrated within a defined austral summer window, approximately November to April, with near-zero suitability during winter. This seasonal restriction confirms that favourable conditions for *Ae. albopictus*-mediated transmission in Mauritius are temporally bounded rather than sustained year-round, consistent with comparable seasonal dynamics reported in Atlantic archipelagos at similar latitudes (33), and with broader evidence that suitability across island settings is structured by seasonal temperature and humidity cycles (4). Relative to the *Ae. aegypti*-based estimates of (9), which peaked in March, our *Ae. albopictus*-parameterised estimates peaked approximately one month later in April, consistent with differences in thermal performance curves of the two species (14). The sustained suitability observed between March and April, despite modest temperature declines (0.5-0.9°C), reflected the persistence of elevated relative humidity during this period. The sharper temperature decline between April and May (1.5-1.8°C), combined with falling humidity, marked the transition to winter and the decline in suitability. Recorded dengue outbreaks in Mauritius, including sporadic transmission events since 2009 and the 2024 epidemic, have consistently occurred within this estimated suitability window; while multiple non-climatic factors drive outbreak occurrence, including importation timing, population immunity and vector density, this temporal concordance lends face validity to the model and is consistent with climate suitability representing a necessary condition for local *Ae. albopictus*-mediated transmission.

The most notable finding was the divergence in suitability profiles between the two viruses. Chikungunya exhibited consistently higher Index P values, broader spatial coverage and a longer seasonal window than dengue: median suitability persisted above the indicative threshold) into May, while dengue rarely exceeded this threshold in any district. Spatially, chikungunya peak suitability was broadly distributed across coastal regions, whereas dengue peak suitability was concentrated in a few western localities, primarily within Black River district. This divergence is attributable to differences EIP thermal sensitivity. The EIP for dengue in *Ae. albopictus* is longer and more temperature-sensitive than for chikungunya, meaning dengue requires warmer conditions to achieve equivalent transmission potential (13,22). Experimental evidence from an altitude gradient study (149-1209m) directly supports this: (54) demonstrated that chikungunya remained transmissible at 20°C, while dengue was efficiently transmitted only at higher temperatures, approximately 28°C. Mauritius’s volcanic topography appears sufficient to differentially constrain dengue suitability while permitting broader chikungunya suitability.

The broader chikungunya suitability estimated here is biologically plausible in the Mauritian context. Genetic studies of the 2005–2006 Indian Ocean epidemic identified the emergence of the E1-A226V mutation, subsequently shown experimentally to enhance the ECSA chikungunya virus fitness in *Ae. albopictus* (55,56). A recent Mauritius preprint further reported an ECSA lineage carrying *Ae. albopictus*-adaptive mutations during the 2025 chikungunya outbreak, closely associated with rainfall-driven increases in vector abundance (57). Because Index P was estimated using temperature and humidity dependent priors alone, this rainfall-related finding highlights additional ecological and entomological drivers of outbreak occurrence beyond transmission suitability. Together, these findings suggest that dengue and chikungunya preparedness in Mauritius should not be treated as interchangeable: the geographic and temporal risk profiles differ, and surveillance and vector control strategies may need to account for these distinctions.

For a non-endemic island setting, climate suitability defines the seasonal window within which virus importation is most likely to trigger local transmission. Importation events during November to April would encounter conditions conducive to onward transmission by *Ae. albopictus*, while introductions during winter would be less likely to sustain transmission chains. Aligning entomological surveillance, early warning systems and vector control activities with the onset of the suitability window, particularly in higher-risk coastal localities, represents a practical application of these findings for public health planning in Mauritius.

Several limitations should be noted. *Ae. albopictus* parameter estimates were sourced primarily from studies conducted outside Mauritius, particularly Reunion Island; while the two islands are ecologically comparable, local entomological variation may not be fully captured. The MVSE framework incorporates temperature and relative humidity but does not model their interactive effects explicitly (44), which may influence estimates in settings with pronounced humidity gradients. The model captures adult-stage climatic effects but does not represent aquatic-stage dynamics, so estimates reflect adult-stage transmission suitability rather than habitat suitability. ERA5-Land reanalysis dataset at approximately 11 km resolution may not capture fine-scale microclimatic variation, particularly in areas with steep elevation gradients. Finally, Index P represents relative climate suitability rather than outbreak probability, an, and virus-specific differences in Index P estimates should be interpreted cautiously given the vector-competence parameterisation used; realised transmission also depends on virus importation, viral genotype, population immunity, vector density and public health response capacity, none of which are captured by the framework.

## Conclusion

This study provides an essential *Ae. albopictus*-parameterised estimates of climate-driven transmission suitability for dengue and chikungunya in Mauritius. Suitability is seasonally restricted to the austral summer for both viruses, but their spatial and temporal profiles diverge: chikungunya suitability is broader, more sustained and less geographically constrained than dengue, reflecting differential thermal sensitivity of the extrinsic incubation period. For a non-endemic island where outbreaks depend on the coincidence of virus importation and suitable conditions, these estimates provide an evidence base for aligning surveillance and vector control with periods and locations of elevated risk. Validation against spatially resolved case data, when available, would test whether district-level suitability estimates correspond to observed transmission intensity. Coupling Index P with importation pressure models based on regional connectivity and traveller volumes could further refine risk windows. Integration of these suitability profiles into existing Mauritian decision-support tools such as ALBOMAURICE could extend their scope beyond vector distribution mapping to include virus-specific transmission risk assessment. This framework is readily transferable to other SIDS where *Ae. albopictus* is a principal vector, providing a template for anticipatory climate-based arbovirus risk stratification.

## Supporting information

Supplementary materials Index P Mauritius

Supplementary_Table_S4_Spatial_IndexP_values

Supplementary_Table_S5_Seasonal_Monthly_IndexP_values

## Data Availability

All data produced in the present study are available upon reasonable request to the authors

## Acknowledgement

Faculty of Health, Medicine and Behavioural Sciences, University of Queensland

1. Operational Research and Decision Support for Prevention, Control and Elimination of Infectious Diseases, Centre for Clinical Research
2. The Ministry of Health and Wellness, Mauritius
3. Cartography Section, Ministry of Housing and Lands, Mauritius
4. This research was supported by an Australian Government Research Training Program (RTP) Scholarship.

## Declaration of generative AI and AI-assisted technologies in the manuscript preparation process

During the preparation of this work, the authors used ChatGPT to revise and refine sections of Google Earth Engine scripts and R code for data extraction and analysis. These were subsequently tested and implemented within the analysis workflow. The authors also used Claude to improve language clarity and readability. After using these tools, the authors reviewed and edited all content as needed and take full responsibility for the content of the published article.

