## Supplementary materials Index P Mauritius for "Divergent climate suitability profiles for dengue and chikungunya transmission by Aedes albopictus in Mauritius"

Corresponding author: Mohabeer Teeluck

Authors: Mohabeer Teeluck, Emma S. McBryde, Oyelola A. Adegboye, Stephan Karl, Benn Sartorius, Eloise B. Skinner

### Supplementary Methods S1: Meteorological data extraction

Daily mean temperature and dewpoint temperature were extracted at the locality level from ERA5-Land reanalysis datasets via the Google Earth Engine (GEE) Data Catalog (Google Earth Engine, 2025) (Supplementary Table S1). Although ERA5-Land reanalysis datasets report a nominal pixel size of ~11 km, locality-level values were extracted using zonal reduction at a finer nominal scale (~5 km). Additionally, in order to avoid missing values, a 5 km buffer was used as a fallback to ensure complete locality coverage. This procedure was applied for extraction completeness only and did not alter the native spatial resolution of the ERA5-Land dataset. Daily values were aggregated to monthly means for the period December 2013 to October 2024.

#### Supplementary Methods S2: Derivation of relative humidity

Relative humidity was estimated from monthly mean temperature and monthly mean dewpoint temperature using the Tetens approximation (Brown et al., 2023). Saturation vapour pressure was first calculated using the Clausius-Clapeyron equation (Alduchov & Eskridge, 1996):

$$e(T) = 6.112 \times \exp\{17.67 \times T / (T + 243.5)\}$$

where $T$ is the monthly mean temperature in °C. The same formula was applied using $T$d (monthly mean dewpoint temperature) to obtain e($T$d). Relative humidity (RH, %) was then computed as:

$$RH = 100 \times e(Td) / e(T)$$ (Lawrence, 2005)

#### Supplementary Methods S3: Index P equations

Index P for each virus, locality ($d$) and time point ($t$) was calculated using the base equation embedded in the MVSE framework (Obolski et al., 2019):

$$\textbf{\textit{Index }} \boldsymbol{P_{virus}(d,t)} = \frac{\boldsymbol{b(d,t)^2\, \phi^{v\to h}(t)\, \phi^{h\to v}\, \gamma_{v,virus}(t)\, \gamma_{h,virus}}}{\boldsymbol{\mu_v(d,t)\,(\sigma_{h,virus} + \mu_h)\,(\gamma_{h,virus} + \mu_h)\,\left(\gamma_{v,virus}(t) + \mu_v(d,t)\right)}}$$

**(1)**

Index P was further adjusted by applying a virus-specific vector competence coefficient as a transmission efficiency multiplier:

$$\textbf{\textit{Index }} \boldsymbol{P^{*}_{virus}(d,t) = vc_{virus} \times P_{virus}(d,t)}$$

**(2)**

where Index P*virus(d,t)* is the vector-competence-adjusted Index P, representing climate-based transmission suitability scaled by the empirically estimated transmission efficiency of *Aedes albopictus* for each virus. Vector competence values (Supplementary Table S3) were applied as a single multiplier following Tegar et al. (2026), who defined vector competence as the combined probability of mosquito-to-host and host-to-mosquito transmission.

For each locality-month combination, biological parameters were repeatedly drawn from their specified prior distributions (Supplementary Table S3), and Index P was recalculated from these sampled values. Uncertainty in Index P was summarised using the 2.5th and 97.5th percentiles of the resulting simulated Index P distribution, consistent with Monte Carlo uncertainty propagation approaches (Stoudt et al., 2021). Index P was estimated for dengue and chikungunya at each locality and month using the R implementation adapted from the Nakase et al. (2023) tutorial (Nakase, 2022/2023).

**Supplementary Table S1**: *Sources of data, spatial scale and temporal resolution for variables.*

| Indicator | Sources | Spatial resolution | Temporal resolution (12/2013 – 10/2024) |
|---|---|---|---|
| Minimum temperature (ºC) | ERA5-Land Daily Aggregated - ECMWF Climate Reanalysis (temperature_2m)<br>Earth Engine Data Catalog<br>(Google Earth Engine, 2025) | ~ 11 km | daily |
| Maximum temperature (ºC) | ERA5-Land Daily Aggregated - ECMWF Climate Reanalysis (temperature_2m)<br>Earth Engine Data Catalog<br>(Google Earth Engine, 2025) | ~ 11 km | daily |
| Mean temperature (ºC) | ERA5-Land Daily Aggregated - ECMWF Climate Reanalysis (temperature_2m)<br>Earth Engine Data Catalog<br>(Google Earth Engine, 2025) | ~ 11 km | daily |
| Dew temperature (ºC) | ERA5-Land Hourly - ECMWF Climate Reanalysis (dewpoint_temperature_2m)<br>Earth Engine Data Catalog<br>(Google Earth Engine, 2025) | ~ 11 km | daily |
| Relative humidity (%) | Calculated using mean temperature and dew temperature<br>(Google Earth Engine, 2025) | ~ 11 km | daily |

**Supplementary Table S2**: *Descriptions and probability distributions of the biological parameters used in the estimation of Index P for DENV and CHIKV transmission by Aedes albopictus. SD denotes standard deviation. Virus-specific transmission efficiency was applied as a fixed, unitless vector competence multiplier.*

| Symbol | Parameter | Type | Value (Mean ± SD) | Distribution | Units | Source |
|---|---|---|---|---|---|---|
| $1/\mu_v$ *(d,t)* | Adult *Aedes albopictus* lifespan | Field-informed prior | (17.71±4.39) Field temp: 16.6-29.2 °C | Normal | days | (Lacroix et al., 2009) |
| *b (d,t)* | Adult *Aedes albopictus* biting rate | Deterministic temperature-dependent equation (Brière equation) | (0.23±0.04) Lab temp: 10.25-38.32 °C | Normal | $\text{bites.mosq.}^{-1}\text{.day}^{-1}$ | (Mordecai et al., 2017) |
| $1/\gamma_{v,DENV}(d,t)$ | Extrinsic *Aedes albopictus*-DENV incubation period | Deterministic temperature-dependent equation (Brière equation) | (7.01±1.51) Lab temp: 10.39-43.05 °C | Normal | days | (Mordecai et al., 2017) |
| $1/\gamma_{v,CHIKV}(d,t)$ | Extrinsic *Aedes albopictus*-CHIK incubation period | Climate derived-temperature dependent functions | (2.78±0.49) Lab temp: 9.62-36.23 °C | Lognormal | days | (Tegar et al., 2026) |
| $1/\gamma_{h,DENV}$ | Intrinsic human – DENV incubation period | Host intrinsic biological parameters | (5.90±1.79) | Gamma | days | (Chan & Johansson, 2012) |
| $1/\gamma_{h,CHIKV}$ | Intrinsic human – CHIK incubation period | Host intrinsic biological parameters | (3.0±1.3) | Lognormal | days | (Boëlle et al., 2008) |
| $1/\sigma_{h,DENV}$ | Human – DENV infectious period | Host intrinsic clinical parameters | (4.0±0.51) | Normal | days | (Duong et al., 2015; Nakase et al., 2023; Nguyen et al., 2013; Nishiura & Halstead, 2007) |
| $1/\sigma_{h,CHIKV}$ | Human – CHIK infectious period | Host intrinsic biological parameters | (5.0±1.0) | Normal | days | (Johansson et al., 2014) |
| $VC_{DENV}$ | Transmission efficiency | Vector competence | 0.354 | fixed value | none | (Hafsia et al., 2025) |
| $VC_{CHIKV}$ | Transmission efficiency | Vector competence | 0.625 | fixed value | none | (Hafsia et al., 2025) |
| $\phi^{v\to h}$ *(t)* | Transmission probability | (Vector to human) temperature-dependent | none | none | none | (Nakase et al., 2023) |
| $\phi^{h\to v}$ *(t)* | Transmission probability | (Human to vector) temperature-dependent temperature-dependent | none | none | none | (Nakase et al., 2023) |

The (d,t) notation indicates that these parameters vary by locality and time through local temperature and humidity. Biting-rate and incubation-period priors were estimated from temperature equations before being used in MVSE.

Supplementary Table S3 *Descriptive summary of climate variables (mean temperature and relative humidity) at National and Districts level. Study period: December 2013 – October 2024*

| **Admin level** | **Mean Temperature (°C)** | | **Relative Humidity (%)** | |
|---|---|---|---|---|
| **National** | **Mean (SD)** | **Median (Range)** | **Mean (SD)** | **Median (Range)** |
| Mauritius | 23.54 (2.11) | 23.72 (17.39-28.76) | 75.52 (7.79) | 76.02 (47.34-92.98) |
| **District** | **Mean (SD)** | **Median (Range)** | **Mean (SD)** | **Median (Range)** |
| Riviere du Rempart | 24.13 (1.94) | 24.28 (19.76-28.28) | 75.16 (7.65) | 75.48 (50.92-91.29) |
| Pamplemousses | 24.02 (1.97) | 24.18 (18.23-28.28) | 75.05 (7.64) | 75.34 (50.83-91.23) |
| Savanne | 23.76 (2.09) | 23.92 (17.93-28.59) | 75.6 (7.74) | 76.23 (47.91-92.22) |
| Flacq | 23.60 (2.03) | 23.78 (18.17-28.26) | 76.61 (7.63) | 77.28 (51.14-92.25) |
| Grand Port | 23.29 (2.11) | 23.47 (17.95-28.06) | 77.54 (7.59) | 78.45 (51.98-92.98) |
| Black River | 23.86 (2.14) | 24.07 (17.4-28.76) | 72.76 (7.95) | 72.7 (47.34-91.29) |
| Port Louis | 23.60 (2.04) | 23.8 (18.68-28.28) | 74.7 (7.66) | 75.03 (49.53-90.84) |
| Moka | 22.98 (2.12) | 23.2 (17.39-27.67) | 75.87 (7.66) | 76.31 (50.53-92.02) |
| Plaines Wilhems | 22.88 (2.18) | 23.12 (17.4-27.92) | 74.66 (7.78) | 74.97 (49.46-92.00) |

### Supplementary Figure S1

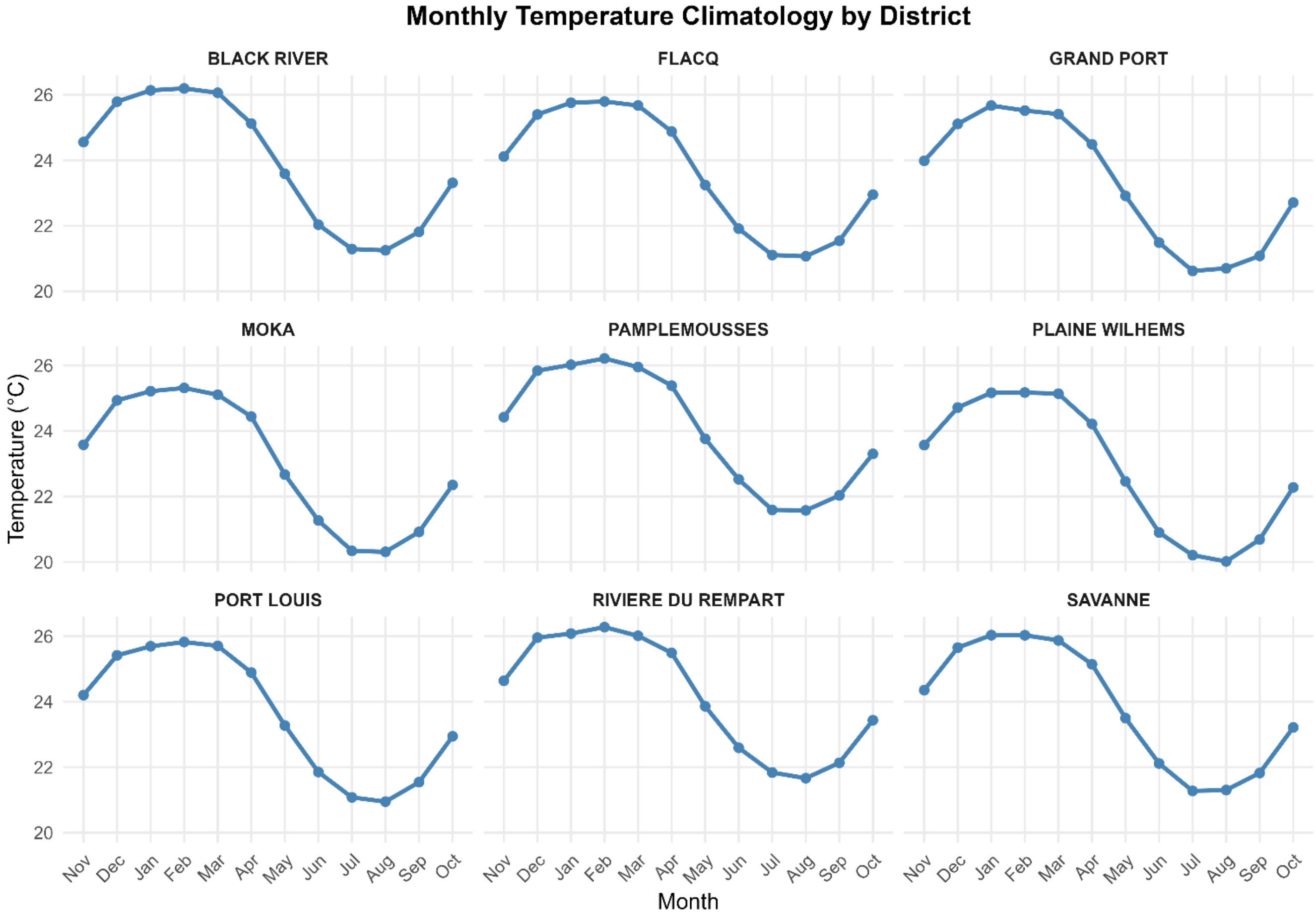


***Figure S1: Monthly temperature climatology across districts in Mauritius, 2014–2024.***

*Lines show district-level monthly median mean temperature derived from locality-level monthly climate summaries. Months are ordered from November to October to display the warm–humid season followed by the cooler winter period. Temperature peaked during the austral summer months and declined during winter, reflecting the expected seasonal thermal pattern across Mauritius.*

### Supplementary Figure S2

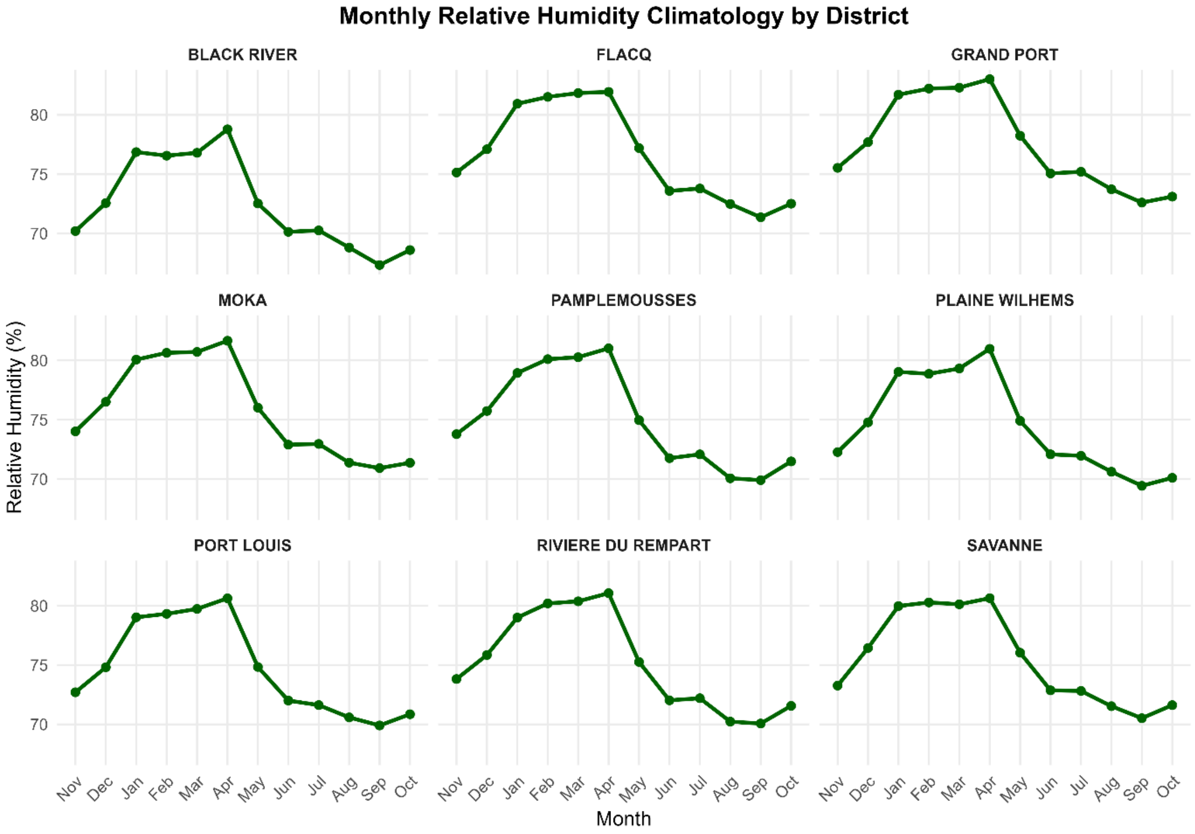


***Figure S2. Monthly relative humidity climatology across districts in Mauritius, 2014–2024.***

*Lines show district-level monthly median relative humidity derived from locality-level monthly climate summaries. Relative humidity was estimated from monthly mean temperature and monthly mean dew-point temperature using the Tetens equation. Months are ordered from November to October to display the warm–humid season followed by the cooler and relatively drier winter period.*

### Supplementary Figure S3

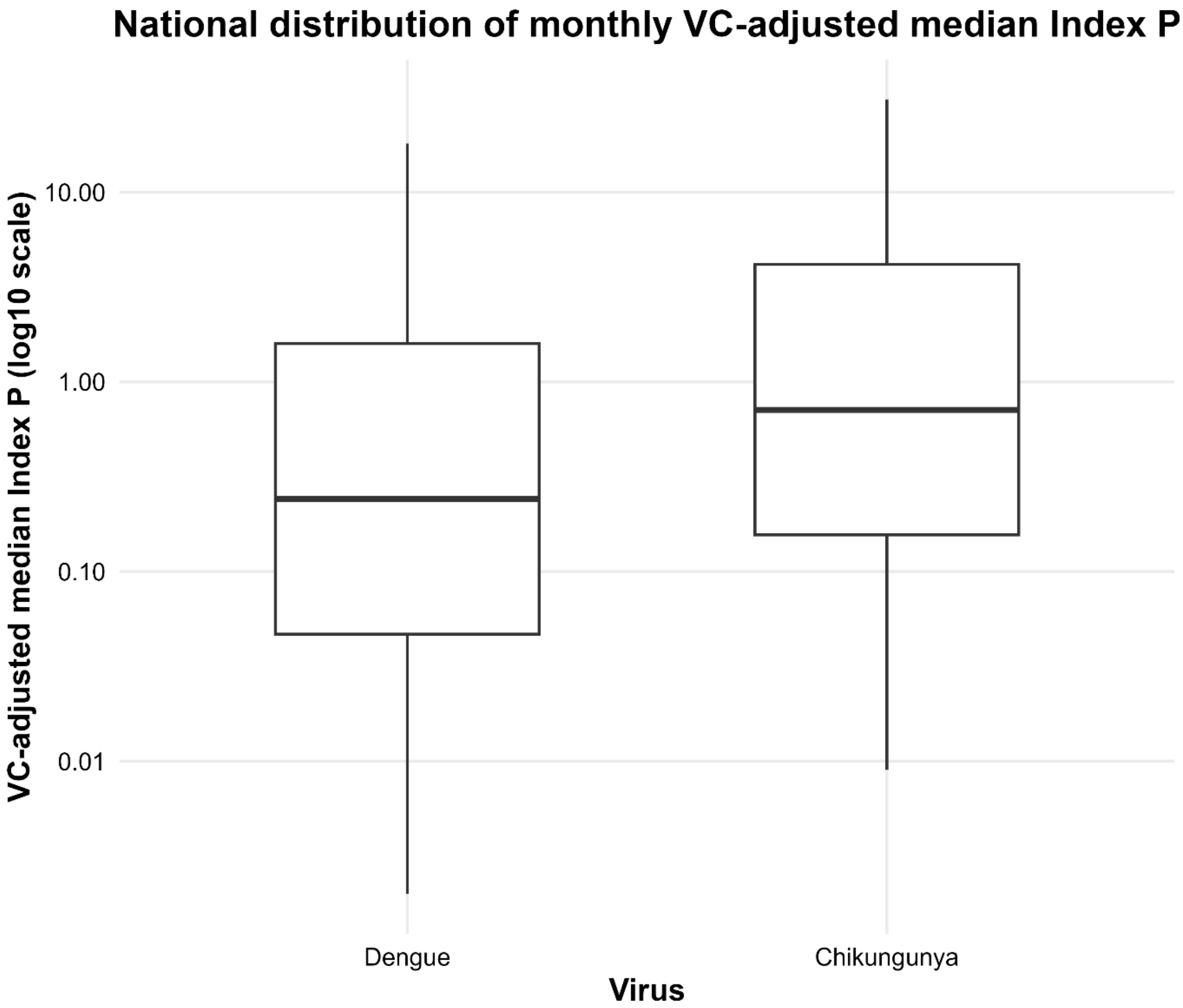


***Supplementary Figure S3. National distribution of monthly VC-adjusted Index P values for dengue and chikungunya across Mauritius, 2014–2024.***

*Boxplots show generated median Index P values across all locality-month observations for each virus. Values are displayed on a log10 scale to improve visualisation of the strongly right-skewed distribution and reduce visual compression caused by high upper-tail values.*

### Supplementary Figure S4.

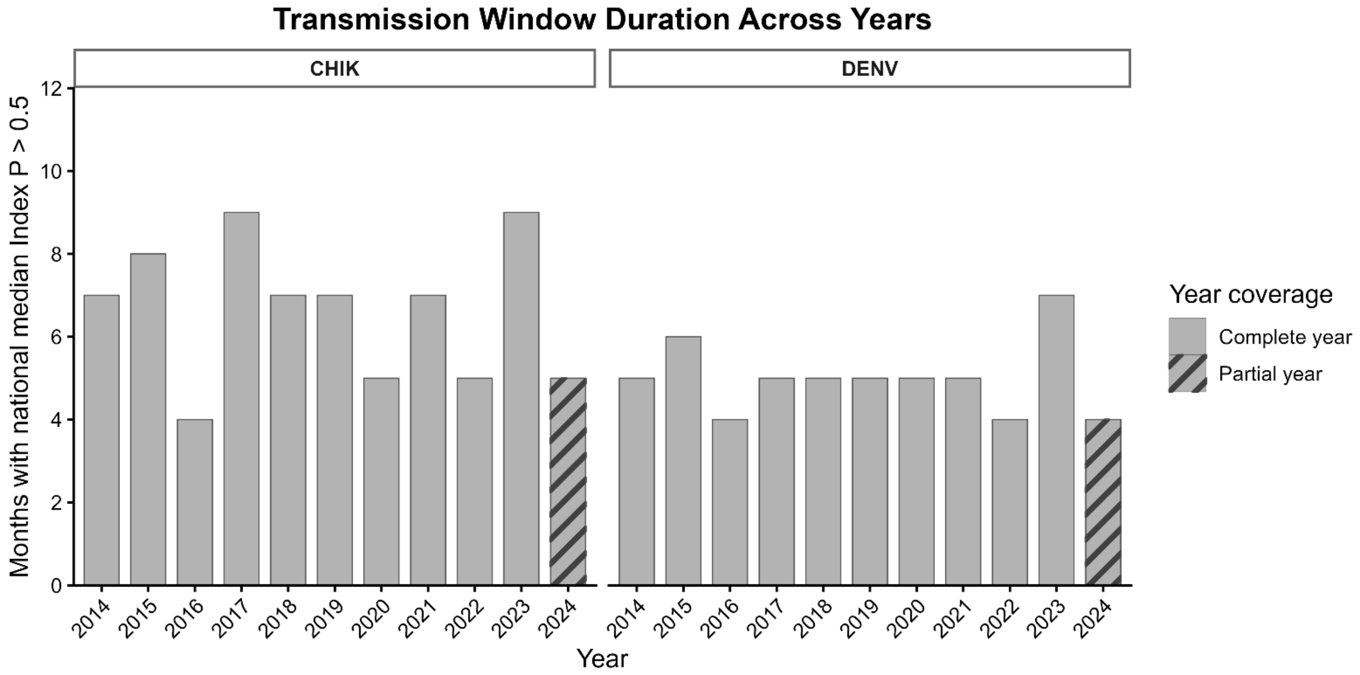


***Figure S4. Annual transmission-window duration for dengue and chikungunya across Mauritius, Jan 2014–Oct 2024.***

*Bars represent the number of months per year with national median VC-adjusted Index P > 0.5, summarised separately for chikungunya and dengue. Hatched bars denote partial-year estimates for 2024, reflecting the availability of data through October only.*
